# OK-AIR study protocol: a longitudinal cluster-randomised 2×2 factorial trial of portable air purification and upper-room UVGI on sick-related absences, indoor air quality, environmental pathogens and social-emotional development in early care and education classrooms (birth–5 years)

**DOI:** 10.64898/2026.03.05.26347562

**Authors:** Changjie Cai, Diane Horm, Barbara Fuhrman, Craig Van Pay, Mingze Zhu, Kristen Shelton, Jason Vogel, Chao Xu

**Author notes:** **Corresponding Authors:** Dr. Changjie Cai, The University of Oklahoma Health Sciences Center, The University of Oklahoma, Oklahoma City, Oklahoma 73104, USA, Dr. Diane Horm, The Early Childhood Education Institute, The University of Oklahoma-Tulsa, Tulsa, Oklahoma 74135, USA, Dr. Barbara Fuhrman, School of Medicine Preventive Medicine and Biostatistics, Uniformed Services University, Bethesda, Maryland 20814, USA.

## Abstract

This protocol is reported in accordance with the SPIRIT 2025 guidelines for clinical trial protocols.

**Introduction:** Young children, from birth to age 5 y are particularly vulnerable to indoor air pollutants and respiratory pathogens. Portable air purifiers (or filtration) and upper-room ultraviolet germicidal irradiation (UVGI) are two widely used interventions with the potential to improve indoor air quality (IAQ) and reduce sick-related absences. However, a review of the literature revealed no real-world randomised studies evaluating their effectiveness in reducing young children’s sick-related absences in early care and education (ECE) classrooms.

**Methods and Analysis:** The OK-AIR study is a longitudinal, cluster-randomised 2×2 factorial trial conducted in Head Start centers using two implementation cohorts: Cohort 1 (five Head Start centers and 20 classrooms from 2023 to 2024) and Cohort 2 (11 centers and 59 classrooms from 2025 to 2026), with expanded inclusion of rural areas. Cohort 1 enrolled 204 children, 48 teachers and 5 site directors, and Cohort 2 enrolled 462 children, 97 teachers and 11 site directors. Within each center, four classrooms are randomised to: (1) control; (2) portable filtration; (3) upper-room ultraviolet germicidal irradiation (UVGI); or (4) both interventions. Cohort 2 was initially planned as a second factorial trial but was amended to a purifier-only design due to funding changes; details are provided in the protocol amendments section. We collect continuous IAQ data, including particulate matter (PM) with aerodynamic diameters ≤1 µm (PM_1_), ≤2.5 µm (PM_2.5_), ≤4 µm (PM_4_), and ≤10 µm (PM_10_); total volatile organic compounds (TVOCs) index; nitrogen oxides (NO_x_) index; carbon monoxide (CO), noise; temperature; and relative humidity, alongside daily child absences. Seasonal environmental surface swabs (dining tables and toilet flooring) are tested by Reverse-Transcriptase quantitative Polymerase Chain Reaction (RT-qPCR) for *Influenza A/B*, *Respiratory Syncytial Virus (RSV)*, *Human Parainfluenza Virus Type 3 (HPIV3)*, *Severe Acute Respiratory Syndrome Coronavirus 2 (SARS-CoV-2)*, and *Norovirus*. IAQ monitoring is structured across Winter, Spring, Summer, and Fall, including designated baseline/off-period weeks to characterize temporal and seasonal variability in environmental measures across classrooms and centers. Multi-informant surveys (Director, Teacher, Parent) capture contextual factors, and children’s social-emotional development is assessed using teacher ratings on the Devereux Early Childhood Assessment (DECA). The primary outcome is the sick-related absence rate, analyzed as cumulative absences over the attendance year while accounting for clustering by school and classroom using generalized mixed-effects models. Secondary outcomes include children’s social-emotional ratings, IAQ metrics and pathogen detection rates; analyses of IAQ incorporate time/seasonal structure, and season-stratified absenteeism analyses will be treated as secondary/exploratory refinements. An economic evaluation will estimate incremental intervention costs and cost-effectiveness/cost-benefit (such as cost per sick-related absence day averted).

**Ethics and Dissemination:** This study was approved by the Institutional Review Board (IRB) at the University of Oklahoma. Findings will be shared through peer-reviewed publications; presentations at local, state, and national conferences; research briefs developed for lay and policy audiences; and community briefings prioritizing the participating early childhood programs and communities.

**Disclaimer:** The views expressed are those of the authors and do not reflect the official views of the Uniformed Services University or the United States Department of War.

**Strengths and Limitations of This Study:**

- Real-world longitudinal cluster RCT: The study uses a rigorous longitudinal cluster-randomised 2×2 factorial design in real-world ECE settings.
- Combined interventions: Interventions target both air filtration and disinfection, allowing for combined and comparative evaluation.
- Objective air-quality monitoring: Continuous monitoring of IAQ metrics provides objective and reliable data on environmental change.
- Environmental pathogen surveillance: qPCR on surface swabs yields an objective biological outcome to triangulate with IAQ and absences.
- Comprehensive context and child measures: Multi-method and multi-reporter data collection includes Head Start attendance records, continuous air monitoring, pathogen detection, contextual surveys completed by center directors, teachers, and parents, and standardized social-emotional assessments (DECA) completed by classroom teachers. Head Start program records providing children’s longer-term health data available through Health Insurance Portability and Accountability Act (HIPAA) authorization.
- Clustered/temporal complexity: Seasonal design accounts for variation over time but may introduce complexity in modeling temporal effects.
- Practical Implications: Study findings will have practical implications for Head Start and other ECE programs striving to maximize child attendance with cost effective strategies.

## 1. INTRODUCTION

### 1.1. Children’s Susceptibility, School Exposures and Health Outcomes

Young children are physiologically more vulnerable to air pollution than older children and adults, and can spend substantial time in classrooms where ventilation, outdoor infiltration, and in-room activities shape exposure. A comprehensive critical review of school indoor air quality (IAQ) highlights that poor IAQ is common, that it is driven by ventilation/operations issues, and it is linked with respiratory symptoms and negative impacts on learning and development; conversely, healthier classroom environments are associated with reduced absences and better performance, underscoring the public-health importance of school IAQ remediation (Sadrizadeh et al., 2022). The review calls for integrated mitigation strategies (ventilation, filtration/disinfection) and larger, methodologically rigorous field studies in schools. Systematic evidence focused specifically on indoor fine particles in ECE environments enrolling young children under age 5 y shows wide variability in indoor PM_2.5_ and indoor/outdoor ratios across settings and seasons. A PRISMA-based (Preferred Reporting Items for Systematic Reviews and Meta-Analyses) review of 66 studies found inconsistent methods and heterogeneous results, concluding that stronger statistical designs are needed to quantify how seasonal, meteorological, activity-based, site-based, and ventilation factors drive indoor PM_2.5_ in classrooms (Cooper et al., 2020).

For children 5 years of age and younger, the OK-AIR target population, systematic reviews of ambient (especially traffic-related) pollutants (NO_2_, O_3_, PM_10_/PM_2.5_) document associations of poor IAQ with a range of adverse outcomes, including asthma, respiratory infections, ear infections, and eczema, with critical windows during pregnancy and the first year of life (Spencer-Hwang et al., 2023). Notably, several reported associations occur at pollutant concentrations within current guideline levels, raising concerns that existing standards may not fully protect very young children. In school-based studies, evidence is less consistent for asthma prevalence (physician-diagnosed asthma) than for asthma-related morbidity. For example, in a study of 1,530 children attending Portuguese nursery and primary schools, indoor air pollutant concentrations were not associated with the prevalence of physician-diagnosed asthma, but were associated with measures of respiratory morbidity. For instance, wheezing was associated with higher NO_2_, lower FEV_1_ was associated with higher O_3_ and PM_2.5_, and associations between PM and asthma/wheeze outcomes were stronger among sensitized (atopic) children (Branco et al., 2020). Together, these findings suggest that indoor air pollution in schools may influence symptom severity and lung function in children with atopy, even when it does not clearly predict asthma diagnosis or prevalence.

Multiple lines of evidence link air pollution to cognitive and neurodevelopmental outcomes in childhood. A systematic review (30 studies) found detrimental associations between early-life exposure (prenatal and postnatal) and global intellective functioning as well as attention/executive function, with PM_2.5_, NO_2_, and PAHs most strongly implicated (Castagna et al., 2022). A 2024 systematic review and meta-analysis that pooled estimates from six studies (n ≈4,860) showed an inverse dose-response relationship between PM_2.5_ and IQ, with the largest decrement in Performance IQ (PIQ); findings were consistent across diverse regions and exposure windows (Alter et al., 2024). In addition, a 2023 systematic review/meta-analysis reported that exposure to PM_10_/PM_2.5_ (and to a lesser extent NO₂) is inversely associated with gross motor development in children, extending concerns beyond cognition to motor domains (Parasin et al., 2023). Classroom-level studies connect air quality to academic proficiency. In Salt Lake County (n=156 primary schools), the frequency of peak PM_2.5_ exposures (not just averages) was associated with a higher percentage of third-graders below grade level in math and English Language Arts (ELA), with effects modification by school socioeconomic composition, evidence that episodic indoor/outdoor pollution events can meaningfully affect learning outcomes (Mullen et al., 2020). Early cognitive skills are also shaped by interacting social and environmental factors: using a large U.S. birth cohort, high ambient isophorone (an air-toxics marker) and lower quality home learning environments were independently associated with lower math scores, suggesting additive or compounding pathways from environmental and social risks (Lett et al., 2017).

### 1.2. Absenteeism in Early Education and Head Start Context

Attendance is a critical mechanism linking exposure, infection risk, and child learning and development. Nationally representative Head Start data (FACES 2009; n=2,842) show that more absences, and especially chronic absenteeism, are associated with fewer gains in early math and literacy during the preschool year; excessive absenteeism also attenuates the benefits of high-quality ECE experiences. These findings position absenteeism as a sensitive, policy-relevant outcome in ECE (Ansari & Purtell, 2018). The broader ECE literature indicates center-based care (including Head Start) supports school readiness, particularly for children facing multiple vulnerabilities, highlighting the importance of maintaining attendance and high-quality environments in federally funded ECE programs (Schochet et al., 2020). The most recent intervention evidence further suggests that absenteeism is modifiable, although effects vary meaningfully by context. A meta-analysis synthesizing 13 randomised and quasi-experimental intervention studies published between January 2020 and March 2025 found a statistically significant reduction in absenteeism favoring intervention group (Li et al., 2026) Study setting was the only significant moderator, with stronger effects observed in single-school implementations, underscoring the importance of local context and implementation conditions for achieving measurable attendance improvements (Li et al., 2026). Because many absenteeism measures aggregate absences across reasons, rigorous trials that isolate sick-related absences and test upstream, environmental strategies to reduce infectious exposure (e.g., improvements in IAQ) are needed to clarify how health-protective interventions translate into attendance benefits in ECE settings.

Portable high-efficiency particulate air (HEPA) purifiers (stand-alone air cleaners) are increasingly used in schools as an engineering control to improve classroom IAQ. For example, in a large block-randomised crossover trial in 17 Los Angeles Unified School District elementary schools, adding portable HEPA air cleaners to classrooms already using MERV-13 HVAC filtration lowered average classroom PM_2.5_ by ∼40% and reduced infiltration of outdoor PM_2.5_ into classrooms compared with non-HEPA conditions (Simona et al., 2025). Evidence linking these exposure reductions to student health outcomes is promising but mixed, which is consistent with the most recent meta-analysis (Li et al., 2026): a pilot school-based intervention among children with asthma achieved significant reductions in classroom PM_2.5_ and black carbon and reported modest evidence of improved lung function (Jhun et al., 2017), whereas a larger factorial cluster-randomised trial across 41 urban elementary schools found no statistically significant reduction in asthma symptom-days with classroom HEPA purifiers, suggesting that clinical benefit may depend on baseline exposures, co-exposures, and implementation fidelity (Phipatanakul et al., 2021). For infection-relevant pathways, classroom experiments conducted during regular instruction show that operating HEPA purifiers can rapidly reduce airborne aerosol concentrations (e.g., >90% reduction within <30 minutes, with ∼5.5 h⁻¹ equivalent air exchange), supporting biologic plausibility for reducing airborne transmission risk in densely occupied rooms (Curtius et al., 2021). Consistent with high filtration performance in other settings, a recent multicenter study reported that HEPA-filtered hospital bathrooms had 95% lower particle number concentrations than bathrooms without HEPA filtration (Cai & Eisenberg, 2024), and a commercial portable air purifier could achieve 99.9% ± 0.1% filtration efficiency using its original HEPA filter when challenged with salt aerosol (Oni et al., 2025). However, direct evidence on airborne viral burden in real-world classrooms remains emerging: a secondary analysis of a placebo-controlled cluster trial measuring 19 respiratory viruses in elementary school classroom air samples detected viruses in nearly all samples and found that HEPA purifiers were not associated with lower odds of “high viral exposure” overall, though viral diversity decreased; importantly, higher measured viral exposure at the school level was associated with higher absenteeism, underscoring the need for well-powered school trials that connect filtration-driven improvements in air quality and pathogen indicators to sick-related attendance endpoints (Sun et al., 2025). To the best of the authors’ knowledge, no real-world randomised trial has evaluated classroom HEPA filtration in early care and education settings serving children aged birth to 5 years, despite this population’s heightened vulnerability.

### 1.3. Current Research Gaps, and Implications for OK-AIR

Taken together, the literature indicates that (1) school and ECE environments are important microenvironments for air-pollution exposure; (2) early exposures are associated with respiratory morbidity, and increasingly with cognitive, executive-function, and motor outcomes; (3) academic proficiency and attendance are sensitive to air-quality conditions (including peaks), and (4) vulnerabilities in Head Start populations make ECE classrooms a high-impact setting for intervention. At the same time, reviews emphasize methodological gaps, limited longitudinal designs, heterogeneous exposure metrics, and few real-world randomised evaluations of engineering controls (filtration/UVGI) in classrooms, especially those enrolling the birth–5 age range (Sadrizadeh et al., 2022).

The OK-AIR study was designed to address these gaps by (1) deploying a cluster-randomised 2×2 factorial design to test portable air purifier (or filtration) and UVGI in real-world ECE classrooms; (2) conducting continuous IAQ monitoring (PM₁, PM₂.₅, PM₄, PM₁₀, TVOCs, NOx, thermal-noise conditions) with a seasonal longitudinal design that captures peaks and transitions; (3) measuring sick-related absences alongside environmental pathogen detection to triangulate exposure-illness-attendance pathways; and (4) situating the trial within Head Start programs, where improving IAQ and reducing illness-related absences can plausibly enhance school readiness and equity in early learning. This integrated approach operationalizes the research priorities repeatedly called for in school IAQ reviews and early childhood health evidence syntheses (Sadrizadeh et al., 2022).

## 2. OBJECTIVES

The primary objective of OK-AIR is to determine whether portable filtration and/or upper-room UVGI reduce sick-related absence rates among young children birth–age 5 in Head Start classrooms. Two main hypotheses are (1) interventions will improve IAQ and reduce pathogen detection, and these changes will correlate with lower absences, and (2) classrooms with filtration and/or UVGI will exhibit lower sick-related absence rates compared to classrooms assigned to the control condition (no intervention); the combined (filtration+UVGI) arm is expected to show the largest effect.

The secondary objectives are to quantify intervention effects on IAQ metrics, and to evaluate whether interventions reduce environmental pathogen detection rates on classroom surfaces. The exploratory objectives are to assess whether changes in IAQ and pathogen burden relate to changes in DECA social-emotional domains, and to explore contextual moderators (e.g., ventilation practices, window use, building inspections, home environment) measured via Director/Teacher/Parent surveys.

## 3. METHODS AND ANALYSIS

### 3.1. Study Design

OK-AIR is a longitudinal (**Figure1a**), cluster-randomised trial implemented in cohorts. Cohort 1 (2023–2024) is a 2×2 factorial design with four parallel classroom-level arms within each Head Start center: control, portable filtration only, upper-room UVGI only, and combined filtration+UVGI (**Figure 1b**). The randomization unit is the classroom (cluster), nested within five Head Start centers (four classrooms per center, and 20 classrooms in total). To characterize temporal and seasonal variability in indoor environmental measures and rates of illness, the study year has been partitioned into Winter, Spring, Summer, and Fall. Within each season (∼3 months), we implement an intervention period and designate rolling baseline/off-period weeks at season boundaries to reduce seasonal confounding (**Figure1a**). The pre-intervention baseline comprises the last week of the prior season plus the first week of the current season, and the post-intervention baseline comprises the last week of the current season plus the first week of the next season. Absence data collection, IAQ monitoring, and environmental sampling continue throughout to support within- and between-season comparisons. Importantly, while this seasonal structure is central for IAQ characterization, the primary absenteeism analysis is prespecified as cumulative over the ECE program year (see Data analysis), with season-stratified absenteeism treated as secondary/exploratory.

**Figure 1a.**
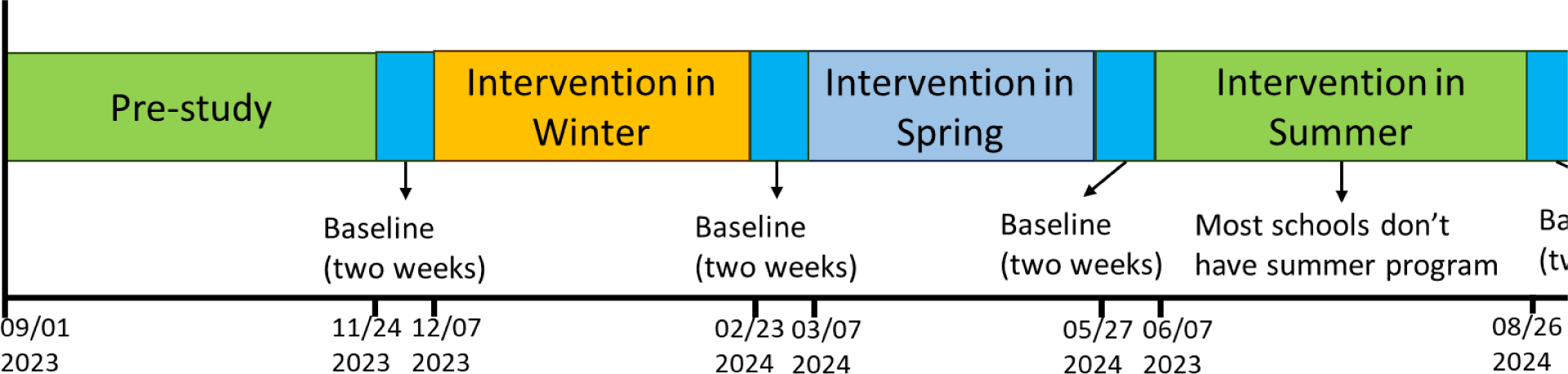
Longitudinal design.

**Figure 1b.**
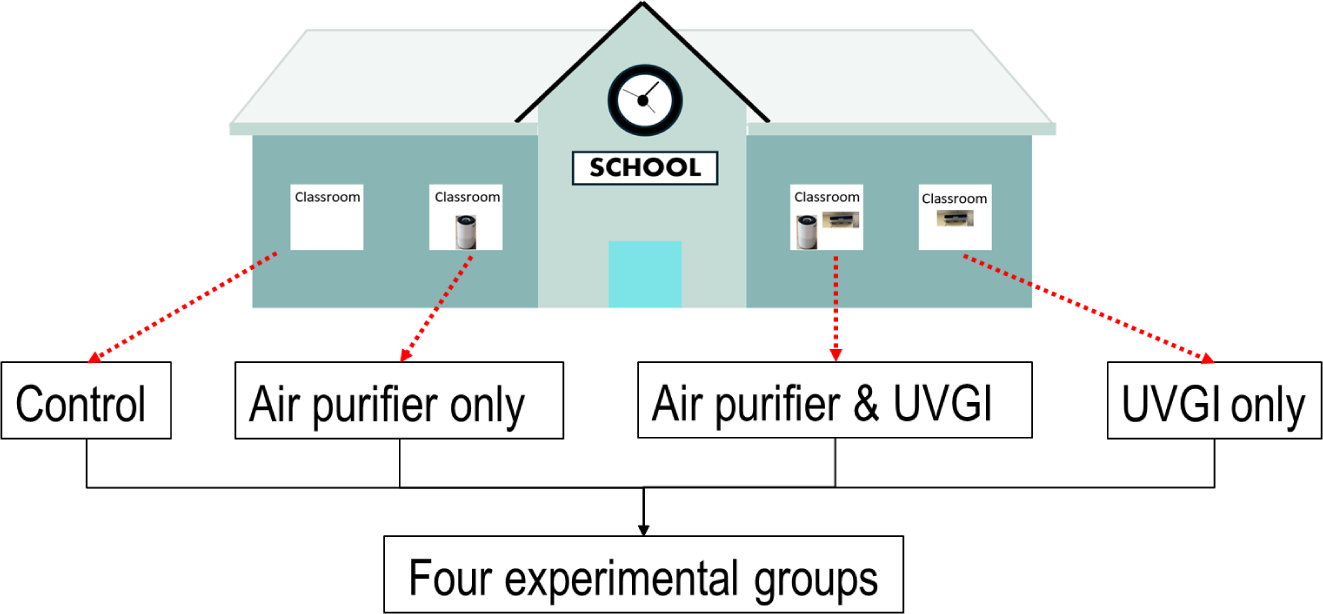
Cluster-randomised 2×2 factorial trial design.

#### 3.1.1. Randomization and allocation concealment

Randomization is conducted at the classroom level. For Cohort 1, each participating Head Start center contributes four classrooms; within each center, classrooms are allocated to the four arms (control, portable filtration only, upper-room UVGI only, and combined portable filtration+UVGI) using a computer-generated random assignment. The allocation list is generated by a designated study team member not involved in day-to-day classroom implementation and is provided to the installation team after baseline procedures are initiated. Because the interventions require visible devices, allocation concealment from classroom staff is not feasible; however, study datasets will use masked arm labels for primary analyses until the analytic dataset is locked.

#### 3.1.2. Blinding

Teachers and classroom staff cannot be blinded to assignment because intervention devices are visible and may affect sound or room layout. To reduce bias, (1) laboratory personnel conducting RT-qPCR analyses will be blinded to intervention assignment using coded sample identifiers, and (2) data analysts will conduct primary analyses using masked arm labels; unmasking will occur after prespecified model code is finalized and the analytic dataset is locked.

This protocol focuses on Cohort 1 (2023–2024), implemented as a longitudinal, cluster-randomised 2×2 factorial trial evaluating two engineering controls (portable filtration and upper-room UVGI) in Head Start classrooms. Cohort 2 (planned 2025–2026) was originally designed as a second cluster-randomised 2×2 factorial trial evaluating portable filtration and an administrative educational program, by following the CDC NIOSH Hazard Hierarchy of Controls. However, Cohort 2 was subsequently amended due to funding changes. The current status and details of protocol amendments are described in Section 3.8.

### 3.2. Sample size and power

Sample size calculations were performed using the National Institutes of Health (NIH) Research Methods Resources power calculator for parallel group-randomised (cluster-randomised) trials (NIH, 2023). We calculated the number of clusters (classrooms) required to achieve the primary aim of reducing children’s absences due to illness under the planned 2×2 factorial cluster-randomised design (four classroom-level arms). Calculations assumed an estimated mean of 12.6 days of absence due to respiratory infection per child-year, an average cluster size of 10 enrolled children per classroom, and an intracluster correlation coefficient of *ICC = 0.03* to account for clustering within classrooms (Murray et al., 1994; Murray & Short, 1995, 1996; Killip et al., 2004; MacIntyre et al., 2015). With a two-sided *α = 0.05* and 80% power, and allowing for 10% attrition, a total sample of 48 classrooms (12 classrooms per arm; approximately 480 children total) was estimated to provide adequate power to detect the independent (main) effects of each intervention and their joint (interaction) effect, assuming standardized effect sizes of *d = 0.30*.

In practice, Cohort 1 implementation was limited to five centers (20 classrooms) due to pragmatic considerations, but classroom rosters were larger than originally assumed. Cohort 1 enrolled 336 children across 20 classrooms (mean cluster size ∼16–17 children/classroom, with some classrooms exceeding 10 children). Because the factorial design estimates main effects by pooling across the two relevant arms (e.g., purifier present vs purifier absent), the effective comparison for each main effect is approximately 10 classrooms versus 10 classrooms, rather than five per arm. Under the original *ICC* assumption (*ICC=0.03*) and average cluster size (∼17), this achieved sample corresponds to a minimum detectable standardized main effect of approximately *d≈0.37* at *α=0.05* and 80% power; therefore, Cohort 1 is powered for small-to-moderate main effects, while the interaction effect will be interpreted as exploratory. We will report achieved cluster sizes and outcome ICC estimates and will conduct sensitivity analyses to assess robustness to unequal cluster sizes, missing data, and alternative ICC assumptions.

### 3.3. Setting and Participants

Cohort 1 is conducted in five Head Start centers in Oklahoma serving children birth–5 years, with four classrooms per center (20 classrooms in total). Only consented children are included in attendance outcomes, and parental consent is obtained for child-level DECA ratings, and survey-linked analyses per Institutional Review Board (IRB) requirements. The Health Insurance Portability and Accountability Act (HIPAA) authorization was obtained from parents for release of their children’s health records collected and maintained by the Head Start program. Classroom inclusion criteria require agreement with intervention installation, IAQ monitoring, absence tracking, and environmental sampling. Classrooms are excluded if physical constraints prevent safe/feasible installation (e.g., ceiling height or layout constraints for upper-room UVGI). **Table 1** summarizes baseline demographic characteristics of participating children. We obtained a total of 204 children’s parents consented in Cohort 1.

**Table 1.** Demographic characteristics of children: Means and Standard Deviations (M ± SD) or Counts and Percentages [N (%)].

| Characteristics | Total (n= 204) | Rural (n= 93) | Urban (n=111) |
| --- | --- | --- | --- |
| Age in month (mean, SD) | 36.67 ± 10.58 | 40.26 ± 9.58 | 33.66 ± 10.49 |
| Gender, N (%) |  |  |  |
| Male | 118 (57.84) | 48 (51.61) | 70 (63.06) |
| Female | 86 (42.16) | 45 (48.39) | 41 (36.94) |
| Race, N (%) |  |  |  |
| American Indian or Alaska Native | 16 (7.84) | 15 (16.13) | 1 (0.90) |
| Asian | 3 (1.47) | 1 (1.08) | 2 (1.80) |
| Black or African American | 36 (17.65) | 10 (10.75) | 26 (23.42) |
| White or Caucasian | 69 (33.82) | 46 (49.46) | 23 (20.72) |
| Bi-Racial/Multiracial | 24 (11.76) | 17 (18.28) | 7 (6.31) |
| Hispanic/Latino | 52 (25.49) | 0 | 52 (46.85) |
| Other | 3 (1.47) | 3 (3.23) | 0 |
| Home language, N (%) |  |  |  |
| English | 162 (79.41) | 82 (88.17) | 80 (72.07) |
| Spanish | 35 (17.16) | 6 (6.45) | 29 (26.13) |
| Other | 1 (0.49) | 4 (4.30) | 2 (1.80) |
| Height in inches (mean, SD) | 33.85 ± 5.86 | 36.81 ± 5.27 | 31.59 ± 5.26 |
| Weight in lbs (mean, SD) | 27.95 ± 9.57 | 32.41 ± 9.38 | 24.53 ± 8.24 |
| BMI (mean, SD) | 16.89 ± 2.28 | 16.58 ± 2.17 | 17.08 ± 2.34 |

### 3.4. Interventions

Cohort 1 includes four arms shown in **Figure 1b**. Portable air purifiers are deployed as stand-alone units equipped with high-efficiency particulate air (HEPA-class) filters. Devices are positioned to maximize room air mixing and coverage without obstructing movement. The number of units and their placement are determined by room volume and manufacturer guidance. Filters are replaced on the recommended schedule (every three months). Upper-room UVGI is provided via wall-mounted fixtures that create an irradiated upper-room zone. All fixtures are installed and commissioned to ensure occupant exposures remain below applicable safety limits, and UVGI lamps are maintained according to manufacturer guidance (replacement annually). Classrooms assigned to the control arm receive no additional filtration or UVGI beyond standard center operations. To support implementation fidelity, installers document device specifications, locations, commissioning results, and all maintenance (including filter and lamp changes). All portable air purifiers are locked so that the children would not accidentally turn them off.

#### 3.4.1. Intervention adherence, fidelity, and co-interventions

Implementation fidelity is assessed through installation and commissioning logs, routine maintenance records (filter and lamp replacements), and periodic classroom checks to confirm that devices are powered on, positioned as installed, and operating at the intended settings during Head Start program hours. Teachers may report any periods when devices were unplugged or turned off (e.g., for noise or classroom activities). Standard center practices (e.g., HVAC operation, window opening, and cleaning routines) are not altered by the study, but some are measured through Director/Teacher surveys and will be considered in sensitivity and contextual analyses.

#### 3.4.2. Safety and adverse event reporting

Potential risks include minor disruption from device noise, trip hazards from cords, and (for UVGI) inadvertent overexposure if fixtures are damaged or mis-positioned. UVGI fixtures are commissioned to maintain exposure levels below applicable safety limits. Study staff maintain an incident log for device problems and any health or safety complaints reported by teachers, parents, or center leadership. Any unanticipated problems involving risks to participants or others will be reported to the IRB in accordance with institutional policy

### 3.5. Data Collection Procedures

#### 3.5.1. Attendance and absence reasons

Teachers record daily attendance and the reason for each absence using a standardized log. Absences are coded as sick-related, not sick, or unknown; sick-related absences are further categorized as respiratory, gastrointestinal, other, or unknown. Attendance logs are reviewed periodically for completeness and internal consistency.

#### 3.5.2. IAQ Monitoring

Each classroom is equipped with a custom, low-cost IAQ monitor that continuously logs particulate matter and environmental parameters. Our first version developed by the University of Iowa, particulate matter (PM₁, PM₂.₅, PM₄, PM₁₀) is measured using a Sensirion SEN-55 environmental node (Stäfa, Switzerland), which integrates an optical particle counter and on-board temperature/relative humidity sensing, and reports VOC Index and NOx Index (dimensionless indices intended for relative trend/event tracking). Carbon monoxide (CO) is measured using a SPEC Sensors 110-102 electrochemical sensor (California, USA). Temperature and relative humidity are also measured using an AHT20 sensor (Aosong/ASAIR, Guangzhou, China). Classroom noise is characterized using a GROK000 module (GROK Lab, Iowa, USA) that records slow time-weighted, A-weighted sound levels (dBA, LAS). Sensor data are queried, time-stamped, and pre-processed by a Teensy 4.1 microcontroller (PJRC, Oregon, USA), stored locally on microSD, and transmitted via a SIM7080G cellular module (SIMCom Wireless Solutions, Shanghai, China; LTE-M/NB-IoT) to a secure server for backup and analysis. For planned analyses, high-frequency data are aggregated to daily and weekly summaries during Head Start program hours, while raw data are retained for sensitivity analyses and QA/QC.

#### 3.5.3. Environmental Surface Sampling and RT-qPCR Viral Analysis

Seasonal surface swabs are collected from dining tables and toilet flooring and processed using Reverse-Transcriptase Quantitative Polymerase Chain Reaction (RT-qPCR) for *Influenza A/B, RSV, HPIV3, SARS-CoV-2*, and *Norovirus (GI/GII/GIV)*. Swabs are stored in DNA/RNA Shield™ buffer (Zymo Research) at −20°C prior to processing. Samples are thawed on ice, clarified by brief centrifugation (16,000 × g for 30 s), and the supernatant is transferred to a clean tube. RNA is extracted using the Quick-DNA/RNA™ MiniPrep Plus Kit (Zymo Research) per manufacturer protocol and analyzed using RT-qPCR with validated primer–probe sets. Remaining RNA is archived at −80°C. Pathogen outcomes include both detection (detected/not detected) and semi-quantitative burden (estimated RNA copy numbers derived from standard curves). Primer/probe sequences and cycling parameters are provided in Supplement.

#### 3.5.4. Contextual Surveys and Acceptability Feedback

We administer three structured surveys to capture center-, classroom-, and household-level contextual factors relevant to indoor environmental quality and child health. The Director Survey captures building operation and ventilation practices, inspection history, humidity management, smoke-free policies, hazardous materials procedures, and staff training/oversight structures. The Teacher Health Survey captures educator characteristics and exposures (including respiratory history and smoking/vaping) and relevant household factors (e.g., housing conditions, ventilation behaviors, and potential indoor sources). The Parent Health Questionnaire captures child health (including respiratory symptoms), attendance barriers, and household exposures and housing conditions. In addition, teachers participate in brief feedback processes (survey and/or focus groups) regarding study participation and the acceptability/feasibility of interventions in operating ECE classrooms. For example, school stakeholders (Head Start directors and classroom teachers/school staff) were involved after Cohort 1 through feedback interviews to improve feasibility and acceptability for Cohort 2. Stakeholders reported that the air-quality monitors were disruptive due to fan noise. In response, we modified the monitors for Cohort 2 by removing the fan and using passive airflow driven by thermal differences to draw air through the device. Stakeholders were not involved in statistical analysis or manuscript preparation. Full instruments are included in the Supplement.

#### 3.5.5. Child Social-Emotional Assessment

Teachers complete age-appropriate DECA instruments following standard program schedules, using the DECA-I/T for infants/toddlers (Mackrain & LeBuffe, 2007) or the DECA-P2 for preschoolers (LeBuffe & Naglieri, 2013). Teachers rate behaviors over the prior four weeks using the standard response scale, and scores are converted to T-scores per instrument guidance. Analyses focus on Total Protective Factors (and domain scores where available), as specified in the Study outcomes section.

#### 3.5.6. Implementation Context: Statewide IAQ Perceptions and Training Needs Survey

As a complementary component to OK-AIR, we conducted a statewide survey of ECE staff in Oklahoma to characterize perceptions of IAQ, self-reported IAQ-related symptoms and concerns, perceived control over classroom environments, and knowledge, confidence, and prior training related to IAQ. The survey was distributed electronically through Oklahoma’s childcare training registry and a partnering Head Start agency and included lead teachers, assistant teachers, and support staff working with children from birth to age 5. Findings from this parallel implementation-focused study will be used to inform interpretation of OK-AIR implementation experiences and to guide development of practical dissemination materials and training resources to support adoption and maintenance of classroom IAQ interventions in ECE settings. This statewide survey is separate from the cluster-randomised trial and is not used to estimate intervention effects on absenteeism, IAQ, or pathogen outcomes.

### 3.6. Data Analysis

Descriptive statistics (means, medians, percentiles, histograms, and time series) are computed for all outcomes by arm and season. For sensor data, we will define school-hours windows and compute daily and weekly aggregates (e.g., mean, 95^th^ percentile, time-above-thresholds metrics). Automated quality checks flag days with low device uptime, missing data, or implausible values. Prior to inferential modeling, we will examine distributional assumptions (residual diagnostics, influential points) and apply transformations as needed.

#### 3.6.1. Primary Analysis for Absenteeism

The primary analysis will test differences by study arm in cumulative sick-related absences over the Head Start program year, accounting for clustering by classroom (and by center as needed). We will model sick-related absence counts using generalized mixed-effects models with offsets for enrolled child-days. A negative binomial link will be used when overdispersion is present, and Poisson models with robust standard errors will be used as a sensitivity analysis. Fixed effects will include indicators for portable filtration assignment, UVGI assignment, and their interaction, allowing estimation of main effects and the combined intervention effect. Random intercepts will be included for classroom (and center/school as needed) to account for clustering of children within classrooms and classrooms within centers.

We will report rate ratios (RRs) and/or odds ratios (ORs) and 95% confidence intervals for the main effects and the interaction. Season-stratified absenteeism analyses will be conducted as secondary/exploratory refinements introduced during implementation to align with the environmental monitoring framework, and these results will be interpreted cautiously and clearly labeled as secondary/exploratory. Sensitivity analyses for absenteeism will include (1) alternative definitions of sick-related absence (e.g., excluding “sick-unknown”), (2) modeling all-cause absences as a robustness check, and (3) excluding days or periods with atypical operation (e.g., closures) if they materially affect denominators.

#### 3.6.2. Secondary Analysis for IAQ and Environmental Pathogens

To estimate intervention effects on IAQ, we will fit linear mixed-effects models using repeated IAQ measures (e.g., daily school-hours aggregates). Fixed effects will include portable filtration assignment, UVGI assignment, and their interaction, along with time, season, and location (urban vs suburban vs rural). Random intercepts will be included for center and classroom to account for clustering and repeated measures. Planned contrasts will estimate main effects (portable filtration present vs absent, UVGI present vs absent) and the interaction (combined vs additive expectation). We will also examine Intervention×Location interactions to explore heterogeneity between urban, suburban and rural settings. Separate models will be fit for each IAQ metric, including PM1, PM2.5, PM4, PM10, temperature, relative humidity, noise, CO and gas indices (TVOC and NOx Index). When IAQ outcomes are skewed or exhibit heteroscedasticity, we will use transformed outcomes or alternative distributions as appropriate. If residual autocorrelation is evident within classroom time series, we will consider correlation structures as sensitivity analyses.

Pathogen outcomes will be analyzed in three complementary ways. First, per-pathogen detection (detected vs not detected) will be modeled using mixed-effects logistic regression, including fixed effects for interventions, season, and location, and random intercepts for classroom (and center as appropriate). Second, classroom-season detection proportions will be analyzed using binomial GLMMs or beta-binomial models to account for extra-binomial variation. A composite outcome (≥1 pathogen detected per round) will also be analyzed using a logistic GLMM. Because multiple-pathogen are tested, pathogen-specific results will be interpreted cautiously as secondary outcomes, and we will emphasize effect sizes and confidence intervals. In addition to binary detection (detected/not detected), we will estimate viral RNA copy numbers for each target using RT-qPCR standard curves and report quantities as copies per swab (and, where surface area is standardized, copies per unit area). Because environmental surface sampling and extraction introduce variable recovery, copy number estimates will be interpreted as relative/semi-quantitative measures of environmental burden rather than absolute exposure dose. For quantitative analyses, copy numbers will be log10-transformed to reduce right-skewness and stabilize variance. We will analyze copy number outcomes using mixed-effects models with fixed effects for intervention assignment (portable filtration, UVGI, and their interaction), season, location (urban/suburban/rural), and surface type (dining table vs toilet flooring), and random intercepts for classroom (and center as appropriate) to account for clustering and repeated measures. Effect estimates will be reported as geometric mean ratios (or differences on the log10 scale) with 95% confidence intervals.

#### 3.6.3. Contextual Analyses for Surveys (Director, Teacher and Parent) and DECA Social-Emotional Outcomes

Survey data will be used to (1) describe baseline contextual conditions at the center, classroom, and household levels, (2) assess baseline comparability across randomised arms, and (3) support covariate adjustment, effect-modification analyses, and sensitivity analyses for the primary and secondary outcomes. Director survey variables will be treated as center-level measures. Teacher survey variables will be treated as classroom/teacher-level measures. Parent survey variables will be treated as child/household-level measures. We will compute descriptive summaries for each survey domain overall and by intervention arm, and will summarize missingness patterns. For multi-item domains where it is meaningful (e.g., counts of housing problems or exposure indicators), we will construct simple summary indices (e.g., number of reported housing issues), and will assess internal consistency when appropriate. Survey-derived variables will be incorporated into outcome models in three main ways, including (1) covariate adjustment using a parsimonious set of prespecified contextual variables with strong conceptual relevance (to avoid over-parameterization given the limited number of clusters), (2) effect modification by testing interaction terms between intervention assignment and selected moderators, and (3) sensitivity analyses comparing results with and without survey-based adjustment and using alternative constructions.

For classroom-season outcomes (IAQ summaries, pathogen detection proportions), parent-level covariates may be included as classroom-aggregated summaries. For child-level outcomes (DECA, where linkable), parent/teacher covariates will be included at the child or classroom level as appropriate, with clustering handled via mixed-effects models. DECA outcomes (Initiative, Self-Regulation, Attachment/Relationships, and Total Protective Factors, as available by age form) will be modeled using linear mixed-effects models, including fixed effects for intervention assignment and relevant covariates (e.g., season and child age/sex when linkable) and random intercepts for classroom (and child when repeated measures are linkable). Ordinal models for DECA categorical bands (Strength/Typical/Need) will be considered as robustness checks. DECA analyses are exploratory and will be interpreted accordingly

#### 3.6.4. Exploratory Mediation and Mechanistic Analyses

To better understand the mediation and mechanistic links, we will explore associations between changes in IAQ and sick-related absence, and between pathogen detection and absence, using two-stage approaches (e.g., regressing classroom/personal/seasonal absence rates on IAQ/pathogen summaries while retaining intervention terms). Formal mediation will be treated as exploratory and interpreted cautiously. We will summarize missingness for each outcome and covariate. For survey covariates with non-trivial missingness, we will consider multiple imputation under a missing-at-random assumption and compare with complete-case analyses.

#### 3.6.5. Complementary Study for Analysis of Statewide IAQ Perceptions and Training Needs Survey

Responses from the statewide ECE staff survey will be analyzed to describe implementation context and training needs relevant to IAQ interventions in ECE settings. Analyses will begin with descriptive statistics (frequencies/percentages for categorical variables; means/medians and dispersion for continuous or ordinal scales) overall and stratified by staff role (e.g., lead teacher, assistant teacher, support staff) and setting. For multi-item domains (e.g., IAQ knowledge/confidence, perceived control, symptom groupings, and training needs), we will construct domain scores using prespecified item groupings and will summarize internal consistency where appropriate. Missing data patterns will be described; analyses will primarily use available data, with sensitivity analyses using multiple imputation for key covariates if missingness is substantial.

To explore correlates of IAQ knowledge, confidence, perceived control, and training needs, we will fit multivariable regression models appropriate to outcome scale (e.g., linear models for continuous scores; ordinal logistic models for ordered responses; logistic models for binary endpoints such as prior IAQ training). Candidate predictors may include staff role, years of experience, program type, geography (urban/suburban/rural where available), and self-reported classroom environmental concerns. These analyses will be exploratory and used to inform dissemination and training strategies rather than to estimate intervention efficacy. Results from the statewide survey will be reported separately and will be used to contextualize the OK-AIR trial’s implementation findings and stakeholder-facing recommendations.

### 3.7. Cost Benefit and Cost Effectiveness Analysis

We will evaluate the interventions from (1) the Head Start center perspective and (2) a limited societal perspective that additionally values parental productivity associated with absences averted. For capital equipment with multi-year life (portable filtration and UVGI fixtures), we will present results for the trial year and a multi-year scenario (e.g., 3–5 years) with 3% annual discounting for costs and benefits; reporting will follow Consolidated Health Economic Evaluation Reporting Standards (CHEERS) 2022 and methods recommended by the Second Panel on Cost-Effectiveness in Health and Medicine (including clear statement of perspective and time horizon) (Husereau et al., 2022). We will use micro-costing from study records and center logs: device purchase, installation/commissioning, filters and UV-C lamps, routine maintenance, electricity (nameplate power × logged runtime × local tariff), cellular data/telemetry, sensor replacement/calibration, and staff time for training/QA. Costs will be expressed in 2024 US dollar (USD) and, where applicable, discounted in multi-year scenarios; all inclusions will be tabulated by perspective in an “impact inventory” as recommended by the Second Panel (Carias et al., 2018).

Cost-benefit analysis (CBA) and cost-effectiveness analysis (CEA) will be conducted. For CBA, we will monetize benefits as (1) sick-related absence days averted (from the primary model) valued using a human-capital wage approach for parental productivity (base case with sensitivity bounds), and (2) where applicable, attendance-linked program revenue. We will additionally consider child health-related benefits that are plausibly related to reduced illness, including healthcare use avoided (e.g., outpatient visits, urgent care/ED visits, medications) if sufficient data are available to support unit cost estimation; otherwise, these will be reported descriptively as part of a cost-consequence framework. For CEA, the principal outcome is cost per sick-related absence day averted. Decision summaries will also report net monetary benefit (NMB) and benefit–cost ratio (BCR), with NMB defined as NMB=λ×Absences Averted−ΔCost across willingness-to-pay (λ) per absence day averted (Stinnett & Mullahy, 1998). Because the interventions may yield meaningful child benefits beyond absenteeism (e.g., improvements in IAQ and reduced environmental pathogen burden), we will also conduct a cost-consequence analysis (CCA) presenting intervention costs alongside key trial outcomes, including the sick-related absences, IAQ metrics (e.g., PM₂.₅), pathogen detection and semi-quantitative burden (copy number), and exploratory DECA outcomes. Therefore, decision-makers can consider a broader set of child-relevant benefits without relying on uncertain monetization assumptions.

Parameter uncertainty will be addressed with probabilistic sensitivity analysis (PSA) using Monte Carlo simulation, and results will be displayed as cost-effectiveness acceptability curves (CEACs) and NMB plots over λ. We will conduct one-way and scenario analyses for key drivers (duty cycle, filter/lamp lifetimes and prices, electricity tariffs, wages, class size). Stratified results by urban vs suburban vs rural location and season will be reported to assess heterogeneity (Briggs et al., 2006).

### 3.8. Modifications to the Protocol and Ongoing Work

OK-AIR was initially planned as two sequential cohorts. Cohort 1 (2023-2024) was implemented as a longitudinal, cluster-randomised 2×2 factorial trial evaluating two engineering controls, including the portable filtration and upper-room UVGI. Cohort 2 (2025-2026) was originally planned as a second cluster-randomised 2×2 factorial trial under an the U.S. Environmental Protection Agency (EPA) funded Children’s Environmental Health Center award (https://cfpub.epa.gov/ncer_abstracts/index.cfm/fuseaction/display.abstractDetail/abstract_id/11420), evaluating one engineering control (portable filtration) and one administrative control (delivery of an educational program). During Cohort 1 implementation, the study team incorporated a structured seasonal monitoring framework (Winter, Spring, Summer, Fall) with designated baseline/off-period weeks to characterize temporal and seasonal variability in indoor environmental measures and to support robust environmental comparisons. While the primary absenteeism estimate remains cumulative sick-related absence over the attendance year, analyses stratified by season were added as secondary/exploratory refinements to align absenteeism interpretation with the environmental monitoring framework. These refinements are described in the Statistical Analysis Plan and will be explicitly labelled as secondary/exploratory in reporting.

On 29 April 2025, the study team received notification from the EPA that the Children’s Environmental Health Center award supporting Cohort 2 would be terminated effective 1 May 2025 due to changes in agency research priorities. The termination was not based on project performance. The study team submitted an appeal, which remains under review at the time of protocol preparation. Due to the unexpected loss of funding, the Cohort 2 implementation plan was amended to retain only the portable filtration intervention (the lowest-cost option for participating schools). In addition, Cohort 2 was expected to expand recruitment to a larger number of rural ECE programs as required by the EPA Center award. However, many rural sites have fewer than four classrooms, which further limits the feasibility of maintaining the original within-center 2×2 factorial allocation structure. Any additional changes to Cohort 2 scope, timing, or analytic plans will be documented with protocol version control and reflected in trial registration and the statistical analysis plan as appropriate.

### 3.9. Ethics and Dissemination

#### 3.9.1. Ethics approval

This study was approved by the Institutional Review Board (IRB) at the University of … (protocol number: 16282). Any required local approvals or agreements were obtained from participating Head Start agencies and centers prior to installation and data collection.

#### 3.9.2. Consent and participant rights

Written informed consent is obtained from parents or legal guardians for child participation. Parents additionally provide HIPAA authorization for release of Head Start health records when applicable. Teachers and site directors provide informed consent for surveys and optional qualitative feedback activities. Given the young age of participants, child assent is not obtained. Participation is voluntary, and families and staff may withdraw at any time without penalty or loss of services.

#### 3.9.3 Confidentiality and data management

Each participant and classroom is assigned a unique study ID. Identifiers and consent documents are stored separately from analytic datasets on secure, access-restricted institutional systems. Sensor data transmitted to the secure server are encrypted in transit where supported and are backed up routinely. Environmental samples are labeled with coded identifiers without personal information. Only authorized study personnel have access to identifiable information. Study results will be reported in aggregate to protect confidentiality. Data will be retained in accordance with institutional policy and applicable agreements.

#### 3.9.4. Trial oversight, monitoring, and interim analyses

Given the minimal-risk nature of classroom environmental interventions, no independent data monitoring committee is planned. The study leadership team conducts ongoing monitoring of implementation fidelity, data completeness, and safety incidents. No formal interim efficacy analyses are planned. Audits may be conducted by the IRB, sponsoring institution, or funders as applicable.

#### 3.9.5. Trial registration, protocol access, and dissemination

The trial is registered on the International Standard Randomised Controlled Trial Number (ISRCTN) (registration #: ; registration date: March 06, 2026). Any substantive protocol amendments will be posted with version control. Findings will be disseminated through peer-reviewed publications and presentations, and through plain-language briefs and community briefings for participating Head Start programs and families. Authorship will follow ICMJE criteria, and results will be reported regardless of the direction or magnitude of effects.

#### 3.9.6. Data sharing and access

After publication of the primary outcomes, the study team will share de-identified datasets, a data dictionary, and analytic code to the extent permitted by the IRB, Head Start data use agreements, and participant consent. Requests for additional materials (e.g., survey instruments, codebooks) will be handled through the corresponding authors.

#### 3.9.7. Funding, sponsor, and competing interests

The Oklahoma Partnership for School Readiness (OPSR) Clearinghouse award (110147) is our original sponsor to support the OK-AIR Cohort 1 study. Cohort 2 planning was supported by a U.S. Environmental Protection Agency (EPA) Children’s Environmental Health Center award (R840630, see Section 3.8); the award was terminated effective 1 May 2025 and an appeal remains under review. The funder reviewed the study proposal and participated in periodic progress meetings. The funder had no role in the study design; data collection, analysis, or interpretation; writing of the report; or the decision to submit the article for publication. The authors declare that they have no known competing financial interests or personal relationships that could have appeared to influence the work reported in this paper.

#### 3.9.8. Ancillary and post-trial care

No ancillary clinical care is provided as part of the trial. Participation does not alter access to routine health services. At study completion, intervention equipment may remain in classrooms or be removed based on agreements with participating centers and available resources; maintenance beyond the study period is not guaranteed by the research team unless specified in agreements.

#### 3.9.9. Trial status

Cohort 1 (2023-2024) has completed enrollment and implementation. Cohort 2 (2025-2026) enrollment and implementation are ongoing as of April 1 2026, contingent on funding and site feasibility as described in Section 3.8.

## 4. STUDY OUTCOMES

### 4.1. Primary Outcome: Absenteeism

The primary endpoint is child absenteeism due to illness, recorded daily by classroom teachers using a standardized attendance log. For analysis, absences are classified as sick-related, non-sick, or unknown. Sick-related absences are further categorized as respiratory, gastrointestinal, other, or unknown. The primary measure is the sick-related absence rate, defined as the number of sick-related absence days divided by the number of enrolled child-days, summarized at the classroom level and child level over the attendance year (and expressed as a rate per 100 child-days). The primary outcome is the cumulative sick-related absence rate over the attendance year, accounting for clustering by classroom (and center where applicable).

### 4.2. Secondary Outcomes: IAQ, Environmental Pathogen Burden, and Contextual Surveys

Continuous IAQ monitoring is conducted in each classroom using a multi-sensor device. Secondary IAQ outcomes include PM₁, PM₂.₅, PM₄ and PM₁₀, as well as CO, temperature, relative humidity, and A-weighted sound level (dBA). The device also reports TVOCs and NOx as dimensionless indices (VOC Index and NOx Index) intended for relative trend/event tracking. IAQ endpoints are pre-specified at the classroom–season level and include (1) school-hours mean PM₂.₅ as the primary IAQ indicator, (2) additional PM metrics (PM₁, PM₄, PM₁₀) and peak metrics (e.g., 95th percentile), (3) time-above-threshold metrics for PM and gas indices; and (4) thermal/noise descriptors (mean temperature, relative humidity, and dBA). IAQ outcomes will also be examined as potential mediators/moderators of intervention effects on absenteeism in exploratory analyses.

In parallel, seasonal environmental surface swabs collected from dining tables and toilet flooring are analyzed by RT-qPCR for Influenza A, Influenza B, Respiratory Syncytial Virus (RSV), Human Parainfluenza Virus Type 3 (HPIV3), Severe Acute Respiratory Syndrome Coronavirus 2 (SARS-CoV-2), and Norovirus (GI/GII/GIV). Pathogen outcomes include (i) pathogen-specific detection (detected/not detected) and (ii) classroom-season detection proportions; a composite outcome (≥1 pathogen detected per sampling round) is also evaluated.

Director, Teacher, and Parent surveys provide secondary contextual measures describing center policies and operations (e.g., ventilation practices, inspection history, humidity management, smoke-free policies), classroom/teacher characteristics (e.g., time in classroom, respiratory health history), and child/household exposures (e.g., housing conditions, crowding, heating/cooking fuels, smoke incursions, pets, pesticide use). Survey variables will be summarized by arm and used as covariates/moderators in secondary and sensitivity analyses.

### 4.3. Exploratory Outcomes: Child Social-Emotional Development

The exploratory outcomes include measures of child social-emotional development. We assess social-emotional competence using the Devereux Early Childhood Assessment (DECA), completed by teachers on the program’s standard schedule. Teachers rate children’s socioemotional skills using the Devereux Early Childhood Assessment for Infants/Toddlers (DECA-I/T; Mackrain & LeBuffe, 2007) or Preschoolers-Second Edition (DECA-P2; LeBuffe & Naglieri, 2013), a widely used, validated set of ratings designed to assess protective and resilience factors in young children using a strengths-based approach. These assessments demonstrate strong internal reliability when completed by teachers, with reported Cronbach’s alpha coefficients of 0.94 for infants and 0.95 for toddlers and preschoolers (LeBuffe & Naglieri, 2012). The DECA-I/T Infant form includes 33 items comprising two scales: Initiative and Attachment/Relationships. The Toddler form (36-items) and the DECA-P2 (35 items) adds a third scale: Self-Regulation. For each version, the individual subscales are aggregated to form the Total Protective Factors score, providing a comprehensive measure of socioemotional competences. Teachers rated the frequency of child behaviors over the previous four weeks using a 5-point Likert scale ranging from 0 (never) to 4 (very frequently). In our study, T-scores for Total Protective Factor scores will be analyzed, with higher scores indicating more positive socioemotional development. For 2-to 5-year-olds, exploratory analyses will also use the Behavioral Concerns subscale scores.

### 4.4. Complementary Outcomes: Statewide IAQ Perceptions and Training Needs Survey

As a complementary component (separate from the cluster-randomised trial), a statewide survey of early childhood education staff characterizes perceived IAQ problems, self-reported IAQ-related symptoms, IAQ knowledge and confidence, perceived control over classroom environmental conditions, and prior training/training needs. These measures will be used to contextualize implementation barriers and facilitators and to inform dissemination and training resources. They are not trial endpoints for estimating intervention effects, but they could potentially guide us for future administrative control strategy, such as delivery of an educational program.

## 5. DISCUSSION

This protocol describes OK-AIR, a longitudinal, cluster-randomised 2×2 factorial trial testing two practical engineering controls, including portable filtration and upper-room UVGI, in Head Start classrooms enrolling children birth through age 5. By focusing on Head Start programs, the study addresses a population with high potential benefit from illness prevention and consistent program attendance. The trial’s design allows estimation of main effects of filtration and UVGI and an assessment of their combined (interaction) effect. In addition to the primary outcome of sick-related absenteeism, OK-AIR integrates continuous IAQ monitoring and environmental pathogen surveillance, as well as multi-informant surveys and child social-emotional assessment (DECA). This study is intended to strengthen causal interpretation and provide practical guidance for schools considering layered air interventions.

### 5.1. Anticipated Contributions

OK-AIR will contribute real-world, randomised evidence on whether portable filtration and UVGI can reduce sick-related absences in ECE classrooms. Absenteeism is a salient, policy-relevant outcome because it affects learning and development opportunities for young children and work disruptions for families. The study will also quantify intervention-associated changes in IAQ (including PM size fractions and time-resolved summary metrics) and in environmental pathogen burden measured by RT-qPCR. Importantly, pathogen outcomes include both detection and semi-quantitative viral RNA burden (estimated copy numbers), which may provide additional sensitivity to changes in environmental contamination beyond simple detected/not detected classification. Finally, the surveys and fidelity/acceptability measures will contextualize implementation (e.g., ventilation practices, building conditions, staff capacity, home exposures) and support exploration of heterogeneity in intervention effects across settings (e.g., urban vs suburban vs rural).

### 5.2. Design Strengths

Several features increase the OK-AIR’s rigor and utility. The factorial, cluster-randomised design in 20 classrooms (5 schools) supports internally valid estimates under real operational constraints. A longitudinal seasonal structure with rolling baselines minimizes seasonal confounding and ensures that comparisons are made within relevant climatic and respiratory-virus contexts. Continuous multi-sensor monitoring provides high-resolution IAQ data, and validated RT-qPCR assays on surface swabs offer objective biological endpoints. The protocol incorporates implementation fidelity metrics (e.g., runtime proxies, maintenance logs) to interpret heterogeneity of effects and to inform feasibility and sustainability. Last but not the least, the Head Start setting enhances equity relevance and the potential for immediate translation to similar centers.

### 5.3. Generalizability

OK-AIR is conducted in Oklahoma Head Start centers, which strengthens relevance for publicly funded ECE programs serving low-income families. Generalizability to other regions will depend on similarities in building stock, ventilation systems, classroom sizes/occupancy, and viral circulation. The interventions themselves, portable filtration and upper-room UVGI, are widely available and scalable interventions, suggesting good potential for broader applicability where installation and maintenance can be supported.

### 5.4. Practical and Policy Implications

If portable filtration and/or UVGI reduce sick-related absences and environmental pathogen detection while improving IAQ, the results will support layered mitigation strategies in ECE settings. Findings can inform procurement and maintenance guidance (e.g., device counts per room volume, filter/lamp replacement schedules) and may help stakeholders prioritize capital investments (e.g., combining filtration with UVGI vs either alone). The study’s implementation and cost-context data (e.g., inspection/training budgets, staffing capacity) will be useful to administrators planning sustainable adoption, and the telemetry-enabled monitoring may inform routine IAQ surveillance in schools.

### 5.5. Limitations and Future Directions

As a field trial in operating ECE classrooms, OK-AIR has several limitations common to pragmatic school-based studies. Blinding of classroom staff is not feasible, introducing potential behavior changes (e.g., window opening, cleaning) once interventions are installed. We mitigate this with standardized guidance, surveys capturing contemporaneous practices, and mixed-effects models that adjust for season and location while accounting for clustering.

Spillover/contamination across classrooms within the same school (e.g., shared hallways) is possible; random effects at the center level and sensitivity analyses at the classroom–season level address shared variance. The study is powered for main effects rather than the interaction; interaction estimates will be interpreted cautiously.

Measurement limitations also warrant consideration. Low-cost sensors can drift and are sensitive to humidity and particle composition; therefore, we include sensor calibration, quality assurance procedures and sensitivity analyses. VOC and NOx “indices” are dimensionless and suited for trend detection rather than absolute concentrations, and analyses will interpret them accordingly. Surface swab RT-qPCR indicates presence of viral RNA on fomites, not airborne concentrations or viability. Copy number estimates are treated as semi-quantitative due to variability in sampling location, swab recovery and extraction efficiency, and analyses will therefore emphasize relative differences and robustness checks. Finally, absence reasons rely on teacher logs and caregiver reports and may be misclassified. We will use standardized categories and include sensitivity analyses (such as all-cause absence and alternative definitions of sick-related absence).

The dataset will support follow-up analyses examining temporal relationships between IAQ dynamics (including peaks and time-above-thresholds metrics), pathogen burden, and absenteeism, including lag structures and potential dose-response patterns. If exploratory analyses signal potential DECA outcomes, future studies with larger sample sizes and longer follow-up could evaluate whether improving indoor environments contributes to broader developmental benefits. Additional implementation research may also identify barriers and facilitators to scale-up, particularly in resource-constrained ECE settings.

### 5.6. Protocol Amendments and Ongoing Work

OK-AIR was originally planned as a two-cohort program of work, with Cohort 1 implemented as a 2×2 factorial trial of two engineering controls (portable filtration and upper-room UVGI) and Cohort 2 initially planned as a second factorial trial evaluating portable filtration alongside an administrative educational program in additional (including more rural schools required by the EPA Children’s Environmental Health Center award) settings. However, an unanticipated funding change required substantial modification of Cohort 2. Specifically, following an EPA notification in April 2025 that the Center award supporting Cohort 2 would be terminated effective May 2025 (not due to project performance), the Cohort 2 plan was amended to prioritize the lowest-cost intervention (portable filtration) and to revise other components contingent on funding and feasibility. These modifications may limit the extent to which Cohort 2 can replicate the factorial design and directly compare combined intervention strategies across a broader set of centers. To preserve transparency and interpretability, all amendments (including the addition of season-stratified absenteeism analyses as secondary/exploratory) are documented through protocol versioning, and future Cohort 2 implementation and analyses will be reported with clear delineation of what was prespecified versus amended.

## Supporting information

Supplemental Table for primer and probe sequences and RT-qPCR thermal cycling conditions for environmental pathogen detection

Supplemental Questionnaires for Center Director, School Teachers/Staff, and Parents

## Data Availability

All data produced in the present study are available upon reasonable request to the authors

## Acknowledgements

We acknowledge that OK-AIR Cohort 1 was supported by the Oklahoma Partnership for School Readiness (OPSR) Clearinghouse, planning for Cohort 2 was supported by the U.S. Environmental Protection Agency (EPA) Children’s Environmental Health Center award (R840630), and implementation of Cohort 2 was supported by Preschool Development Grant funding, secured with assistance from OPSR.

## Competing interests

The authors declare that they have no known competing financial interests or personal relationships that could have appeared to influence the work reported in this paper.

## Author contributions

Conception and design: [CC, DH, BF]

Protocol development and methodology: all authors [CC, DH, BF, CVP, MZ, KS, JV, CX]

Funding acquisition: [CC, DH, JV]

Project administration/coordination: [CC, DH]

Statistical/analysis plan: [CC, DH, BF, CVP, MZ, CX]

Drafting the manuscript: [CC, DH, BF]

Critical revision for important intellectual content: all authors [CC, DH, BF, CVP, MZ, KS, JV, CX]

All authors approved the final manuscript and agree to be accountable for all aspects of the work.

## Notes

### Competing Interest Statement

The authors have declared no competing interest.

### Clinical Trial

ISRCTN78764448

### Author Declarations

The Institutional Review Board (IRB) of the University of Oklahoma gave ethical approval (IRB#:16282) for this work.

