## Supplemental Table for primer and probe sequences and RT-qPCR thermal cycling conditions for environmental pathogen detection for "OK-AIR study protocol: a longitudinal cluster-randomised 2×2 factorial trial of portable air purification and upper-room UVGI on sick-related absences, indoor air quality, environmental pathogens and social-emotional development in early care and education classrooms (birth–5 years)"

**Table S1.** Primer and probe sequences and RT-qPCR thermal cycling conditions for environmental pathogen detection

| Pathogen | Sequences (5' to 3') | | Thermal Cycling Parameters | Reference |
|---|---|---|---|---|
| SARS-CoV-21 | Forward | GACCCCAAAATCAGCGAAAT | 25ºC for 2 min, 50ºC for 15 min, 95ºC for 2min; then 45 cycles of 95ºC for 3 sec and 55ºC for 30 sec | Lu, et al. (2020) |
| | Reverse | TCTGGTTACTGCCAGTTGAATCTG | | |
| | Probe | /VIC/ACCCCGCATTACGTTTGGTGGACC /BHQ1/ | | |
| Influenza A | Forward 1 | CAA GAC CAA TCY TGT CAC CTC TGA C | 25ºC for 2 min, 50ºC for 15 min, 95ºC for 2min; then 45 cycles of 95ºC for 15 sec and 55ºC for 30 sec | CDC (2022) |
| | Forward 2 | CAA GAC CAA TYC TGT CAC CTY TGA C | | |
| | Reverse 1 | GCA TTY TGG ACA AAV CGT CTA CG | | |
| | Reverse 2 | GCA TTT TGG ATA AAG CGT CTA CG | | |
| | Probe | /FAM/TGC AGT CCT /ZEN/ CGC TCA CTG GGC ACG/3IABkFQ/ | | |
| Influenza B | Forward | TCC TCA AYT CAC TCT TCG AGC G | | |
| | Reverse | CGG TGC TCT TGA CCA AAT TGG | | |
| | Probe | /YakYel/CCA ATT CGA/ZEN/ GCA GCT GAA ACT GCG GTG/3IABkFQ/ | | |
| Norovirus | Forward | ATGTTCAGATGGATGAGATTCTC | 50ºC for 20 min, 95ºC for 2min; then 45 cycles of 95ºC for 15 sec and 60ºC for 30 sec | Miura et al., (2013) |
| | Reverse | TGTGAATGAAGATGGCGTCGA | | |
| | Probe | /56-FAM/AGC ACG TGG GAG GGC GAT CG/3BHQ_1/ | | |
| RSV | Forward | CTCCAGAATAYAGGCATGAYTCTCC | | Hughes et al., (2022) |
| | Reverse | GCYCTYCTAATYACWGCTGTAAGAC | | |
| | Probe | /5HEX/ TAA CCA AAT /ZEN/ TAG CAG CAG GAG ATA GAT CAG /3IABkFQ/ | | |
| HPIV3 | Forward | ACC AGG AAA CTA TGC TGC AGA ACG GC | 50ºC for 30 min, 95ºC for 10 min; then 45 cycles of 95ºC for 15 sec, 54ºC for 5 sec, 60ºC for 30 sec | Osiowy (1998) |
| | Reverse | GAT CCA CTG TGT CAC CGC TCA ATA CC | | |
| | Probe | /56-TAMN/GGT ATC CAT CAT GTT TAG GAG CTC T/3BHQ_2/ | | |
