## Supplemental Questionnaires for Center Director, School Teachers/Staff, and Parents for "OK-AIR study protocol: a longitudinal cluster-randomised 2×2 factorial trial of portable air purification and upper-room UVGI on sick-related absences, indoor air quality, environmental pathogens and social-emotional development in early care and education classrooms (birth–5 years)"

**Environmental Questionnaire for Center Directors**

Dear Center Directors,

This survey will ask you to report about your building's physical structure, policies, and procedures that may influence the indoor air quality. This information will be kept confidential and will be used to understand the impact of our air quality intervention.

For some items, you may want to consult with your building/custodial manager. Feel free to have them respond to the topics under their supervision.

**Center Name** ________________________________________

**Director Name** ______________________________________

### VEHICLES NEAR THE BUILDING

1. Does the school have school buses?
   - ○ Yes
   - ○ No

2. Please estimate the number of school buses and family cars that idle near the entrance of your center each day.
   - ○ Number of school buses ___________
   - ○ Number of family cars ___________

3. Please estimate the length of the idle time of the school buses and family cars.
   - ○ School buses: _____hours _______minutes (per day)
   - ○ Family vehicles _____hours _______minutes (per day)
   - ○
   - ○ **WINDOWS**
   - ○ Are classroom windows opened *during* class time?
   - ○ Yes, Daily
   - ○ Yes, Weekly
   - ○ Yes, Biweekly
   - ○ Yes, Monthly
   - ○ No
   - ○
   - ○
   - ○ Are classroom windows opened *outside of* class time?
   - ○ Yes, Daily
   - ○ Yes, Weekly
   - ○ :Yes, Biweekly
   - ○ Yes, Monthly
   - ○ Yes, Other frequency (please specify): __________
   - ○ No

4. During which times of the year are windows opened? *Choose all that apply.*
   - ❑ Winter

- ❑ Spring
- ❑ Summer
- ❑ Fall

**BUILDING INSPECTIONS**

5. Does your center conduct periodic inspections for cracks or leaks in the building foundation, walls, and roof?
   - ○ Yes, Monthly
   - ○ Yes, Quarterly
   - ○ Yes, Bi-annually (Twice a year)
   - ○ Yes, Annually
   - ○ Yes, Other frequency (please specify): ____________
   - ○ No

6. Does your center conduct periodic inspections of the plumbing system?
   - ○ Yes, Monthly
   - ○ Yes, Quarterly
   - ○ Yes, Bi-annually (Twice a year)
   - ○ Yes, Annually
   - ○ Yes, Other frequency (please specify): ____________
   - ○ No

7. Does your center conduct periodic inspections of the heating, ventilation, and air-conditioning system?
   - ○ Yes, Monthly
   - ○ Yes, Quarterly
   - ○ Yes, Bi-annually (Twice a year)
   - ○ Yes, Annually
   - ○ Yes, Other frequency (please specify): ____________
   - ○ No

8. Does your center maintain ASHRAE (American Society of Heating, Refrigerating and Air-Conditioning Engineers) ventilation standards?
   - ○ Yes
   - ○ No
   - ○ I don't know
   - ○ Other standards (please specify): ____________

9. Does your center conduct periodic inspections for condensation in and around the school facilities?
   - ○ Yes, Monthly
   - ○ Yes, Quarterly
   - ○ Yes, Bi-annually (Twice a year)
   - ○ Yes, Annually
   - ○ Yes, Other frequency (please specify): ____________
   - ○ No

10. Does your center conduct periodic inspections for mold?

- ○ Yes, Monthly
- ○ Yes, Quarterly
- ○ Yes, Bi-annually (Twice a year)
- ○ Yes, Annually
- ○ Yes, Other frequency (please specify): ____________
- ○ No

### CONTROLLING THE INDOOR ENVIRONMENT

11. a. Does your center experience problems with high moisture levels (for example, wet spots, sweating pipes, increased pest activity, rotting wood, stained ceiling and walls, visible water leaks, etc.)?
    - ○ Yes
    - ○ No

    b. If yes, does center respond within 48-hours?
    - ○ Yes
    - ○ No

12. a. Does your center have a plan for how to address mold problems?
    - ○ Yes
    - ○ No

    b. If yes, has the mold intervention plan been implemented or acted upon in the past year?
    - ○ Yes
    - ○ No

13. a. Does your center maintain indoor relative humidity below 60%?
    - ○ Yes
    - ○ No, humidity level is measured but not kept below 60%.
    - ○ No, humidity level is not measured.

    b. If yes, how does the school ensure humidity is kept below 60%? *Choose all that apply.*
    - ❑ Dehumidifiers
    - ❑ Regular ventilation
    - ❑ Climate-controlled HVAC system
    - ❑ Other methods (please specify): ____________

14. a. Does your center have a smoke-free school policy?
    - ○ Yes
    - ○ No

    b. If yes, does this policy extend to electronic cigarettes or vaping devices?
    - ○ Yes
    - ○ No

### HAZARDOUS MATERIALS

15. Does your center have a plan for how to use, label, store, and dispose of hazardous materials? ("hazardous materials" refer to substances that can be corrosive, flammable, reactive, or toxic. This may include cleaning chemicals, science experimental chemicals, maintenance supplies, and any other materials potentially harmful to health or the environment.)
    - ○ Yes
    - ○ No

16. Please specify which materials your school commonly uses. *Choose all that apply.*
    - ❑ Cleaning chemicals
    - ❑ Laboratory chemicals
    - ❑ Maintenance supplies (paints, solvents, etc.)
    - ❑ Pesticides or herbicides
    - ❑ Other (please specify): ____________

17. Does your center keep an inventory of hazardous materials, and if so, how frequently is the inventory updated?
    - ○ Yes, updated monthly
    - ○ Yes, updated quarterly
    - ○ Yes, updated bi-annually (twice a year)
    - ○ Yes, updated annually
    - ○ Yes, other frequency (please specify): ____________
    - ○ No

18. a. Does your center have a policy to purchase low-emitting products?
    - ○ Yes
    - ○ No

    b. If yes, which categories of products does this policy cover? *Choose all that apply.*
    - ❑ Cleaning supplies
    - ❑ Paints and coatings
    - ❑ Furnishings (e.g., carpets, furniture)
    - ❑ Building materials
    - ❑ Office supplies (e.g., markers, adhesives)
    - ❑ Toys
    - ❑ Other (please specify): ___________

### ADMINISTRATIVE POLICIES

19. Does your center have a health council that addresses the physical school environment?
    - ○ Yes
    - ○ No

20. Does your center have someone (other than the center director) who oversees custodial, maintenance, and environmental issues?
   - O Yes
   - O No

21. Does your center require training for custodial or maintenance staff on the use, labeling, storage, and disposal of hazardous materials?
   - O Yes
   - O No

22. Does your center require training for custodial or maintenance staff on how to address mold problems?
   - O Yes
   - O No

23. Does your school require training for custodial or maintenance staff on indoor air quality?
   - O Yes
   - O No

*The following items ask about the cost of inspections and training. If you are not the person who oversees the budget for these items, please write the name and contact information for the person we should ask.*

24. What is the center's overall annual cost of building inspections?

    ______________________________________

25. What is the center's overall annual cost of training staff on hazardous materials, mold, and other indoor air quality education?

    ______________________________________

**OK-AIR Study**

**Teacher Questionnaire – Baseline Health Assessment**

**Study ID: ______________________________**

**About the person completing the questionnaire:**

1. **My name: _________________________________**

2. **I work in the following classroom: ___________**

   **Role: ______________________**

   **Hours per week spent in the classroom: _______**

3. **Are you of Hispanic, Latino, or Spanish origin?**
   - ○ Yes
   - ○ No

4. **What is your race(s)?** *Please mark ALL that apply.*
   - ❑ American Indian or Alaska Native
   - ❑ Asian
   - ❑ Black or African American
   - ❑ Native Hawaiian or other Pacific Islander
   - ❑ White or Caucasian
   - ❑ Other (please specify): _____________
   - ❑ I prefer not to say

5. **What is your gender?**
   - ○ Male
   - ○ Female
   - ○ Other: ___________
   - ○ I prefer not to say

6. **In general, would you say that your overall health is:**
   - ○ excellent
   - ○ very good
   - ○ good
   - ○ fair
   - ○ poor

**ASTHMA**

7. **Has a doctor or other health professional ever told you that you had asthma?**

   - ○ Yes
   - ○ No [S*kip to item 11*]

8. **Do you still have asthma?**

   - ○ Yes
   - ○ No [*Skip to item 11*]

9. **During the past 12 months, have you had an episode of asthma or an asthma attack?**

   - ○ Yes
   - ○ No [*Skip to item 11*]

10. **Questions about asthma severity**

    a. **In the past 4 weeks, how much of the time did your asthma keep you from getting as much done at work, school or at home?**
       - ○ All of the time
       - ○ Most of the time
       - ○ Some of the time
       - ○ A little of the time
       - ○ None of the time

    b. **During the past 4 weeks, how often have you had shortness of breath?**

       - ○ More than once a day
       - ○ Once a day
       - ○ 3 to 6 times a week
       - ○ Once or twice a week
       - ○ Not at all

    c. **During the past 4 weeks, how often did your asthma symptoms (wheezing, coughing, shortness of breath, chest tightness or pain) wake you up at night or earlier than usual in the morning?**

       - ○ 4 or more nights a week
       - ○ 2 or 3 nights a week
       - ○ Once a week
       - ○ Once or twice
       - ○ Not at all

    d. **During the past 4 weeks, how often have you used your rescue inhaler or nebulizer medication (such as albuterol)?**
       - ○ 3 or more times per day
       - ○ 1 or 2 times per day
       - ○ 2 or 3 times per week
       - ○ Once a week or less

    e. **How would you rate your asthma control during the past 4 weeks?**
       - ○ Not controlled at all
       - ○ Poorly controlled
       - ○ Somewhat controlled
       - ○ Well controlled
       - ○ Completely controlled

f. **In the past 12 months, how many emergency department visits have you had due to asthma (that did not result in a hospitalization)? ________**

g. **In the past 12 months, how many inpatient hospitalizations have you had due to asthma? ________**

**CHRONIC PULMONARY OBSTRUCTIVE DISEASE (COPD)**

11. **Has a doctor, nurse, or other health professional ever told you that you have COPD, emphysema, or chronic bronchitis?**

    - ○ Yes
    - ○ No

**WEIGHT, DIABETES, & PHYSICAL ACTIVITY**

12. **What is your height? ____________ in**

13. **What is your current weight? ____________ lbs**

14. **What is your highest weight (other than in pregnancy)? ____________ lbs**

15. **During the past 12 months, have you tried to lose or control your weight?**
    - ○ Yes
    - ○ No

16. **Have you EVER been told by a doctor or other health professional that you have any of the following: prediabetes, impaired fasting glucose, impaired glucose tolerance, borderline diabetes, or high blood sugar?**
    - ○ Yes
    - ○ No

17. **Do you take any oral medications to treat diabetes?**
    - ○ Yes
    - ○ No

18. **Do you use injectable insulin or an insulin pump to manage diabetes?**
    - ○ Yes
    - ○ No

19. **How much time do you spend sitting on an average work day?**

    __________ minutes each day

    or __________ hours each day

20. **How much time do you spend weekly doing vigorous activity?**

    __________ minutes each week

    or __________ hours each week

**21. How much time do you spend walking in an average week?**

__________ minutes each week

or __________ hours each week

**22. If you know it, please tell us how many steps you take daily?**

___________ steps daily or

- O I do not use a pedometer

**SMOKING HISTORY**

**23. a. Have you smoked at least 100 cigarettes in your lifetime?**

- O No [*Skip to Item 24*]
- O Yes, and I currently smoke
- O Yes, but I no longer smoke

**b. How old were you when you first started smoking?**

___________ years of age

**c. How old were you when you last quit smoking?**

___________ years of age

**d. In total, how many years have you smoked (please subtract years in which you were not smoking)?**

___________ years

**e. Over this period, what is the average number of cigarettes you smoke(d) per day?**

___________ cigarettes per day

**24. Have you ever used an electronic vaporizer such as an e-cigarette?**

- O Never
- O Once or twice
- O Occasionally but not regularly
- O Regularly in the past
- O Regularly now

**25. During the LAST 30 DAYS, on how many days (if any) have you used electronic cigarettes (e-cigarettes)?**

- O None
- O 1-2 days
- O 3-5 days
- O 6-9 days
- O 10-19 days
- O 20-30 days

**HOUSING**

26. **What is your current address?**
    STREET ADDRESS:___________________________________
    CITY:_______________________
    STATE:________
    ZIP:_____________

27. **How long have you lived at your current address?**

    __ __ Years __ __ Months (*If less than 12 months*)

28. **What is the type of housing where you currently live?**
    - O Apartment
    - O Duplex or townhouse
    - O Single family home
    - O Mobile home
    - O Temporary housing (for example, women's shelter, homeless shelter)
    - O Motel/ hotel
    - O Boat, RV, or van
    - O Other (describe): ___________________________

    a. **This house, apartment, or mobile home is:**
    - O Owned by you or someone in this household with a mortgage or loan (include home equity loans)
    - O Owned by you or someone in this household free and clear (without a mortgage or loan)
    - O Rented
    - O Occupied without payment of rent
    - O Not applicable
    - O Other (describe): __________________________

    b. **How much do you spend on your rent or mortgage per month?** $__________

    c. **How many rooms are there in your current home?** Please include all rooms such as the kitchen and living room, but not bathrooms or hallways. _________

    d. **What is the approximate size of your house in square feet?** _______ sq. ft.

    e. **How many people live in your current home?**
    ________ adults
    ________ children (aged 0-18 y)

    Age of the youngest person living in your home: ________ years _________ months

    f. **In your current home, how many people share your bedroom?**
    - O I have my own bedroom.
    - O I share the bedroom with 1 person.
    - O I share the bedroom with 2 people.
    - O I share the bedroom with more than 2 people.

g. **How is your current home heated?**
- ○ Built-in electric units
- ○ Warm air furnace (central heating unit run on gas, propane, etc.)
- ○ Floor, wall, or pipeless furnace (does not use ducts to move warm air throughout the home)
- ○ Steam or hot water
- ○ Other means: ________________________
- ○ Not heated
- ○ I do not know

h. **What fuel is used most for cooking in your home?**
- ○ Utility gas
- ○ Electricity
- ○ Bottled, tank or LP gas
- ○ Wood
- ○ Fuel oil, kerosene
- ○ Coal or coke

i. **Do you use the following equipment in your home?** *Check all that apply.*
- ❑ Central air conditioning (HVAC)
- ❑ Air conditioning window / wall unit(s)
- ❑ Humidifiers(s)
- ❑ Air filtration unit
- ❑ None of these

j. **Do you use any of the following regularly in your home?** *Check all that apply.*
- ❑ Air fresheners or sprays
- ❑ Scented candles
- ❑ Incense
- ❑ None of these

k. **Do you have pets living indoors in your home?** *Check all that apply.*
- ❑ No
- ❑ Yes, cat(s)
- ❑ Yes, dog(s)
- ❑ Yes, bird(s)
- ❑ Yes, other: ______________________

l. **How often do you open windows in your home for ventilation?**

| **In spring:** | **In summer:** | **In fall:** | **In winter:** |
|---|---|---|---|
| ○ Every day<br>○ At least once most weeks<br>○ Occasionally<br>○ Rarely | ○ Every day<br>○ At least once most weeks<br>○ Occasionally<br>○ Rarely | ○ Every day<br>○ At least once most weeks<br>○ Occasionally<br>○ Rarely | ○ Every day<br>○ At least once most weeks<br>○ Occasionally<br>○ Rarely |

**HOUSING QUALITY**

**29. Overall, how would you describe the physical [or structural] condition of your current home?**
- ○ Excellent
- ○ Good
- ○ Fair
- ○ Poor

**30. Now I am going to ask you some questions about problems that people have in some homes or apartments. How much of a problem is…**

a. **A heating system that does not work?**
- O Big Problem
- O Small Problem
- O No Problem at all

b. **Flooding or water infiltration?**
- O Big Problem
- O Small Problem
- O No Problem at all

c. **Mold or mildew?**
- O Big Problem
- O Small Problem
- O No Problem at all

### ENVIRONMENTAL HEALTH

**31. In the last 12 months, how often has anyone, including visitors, smoked tobacco inside your home?**
- O Daily
- O Weekly
- O Monthly
- O A few times
- O Never

**32. In the last 12 months, how often has tobacco smoke entered your home from somewhere else in or around the building?**
- O Daily
- O Weekly
- O Monthly
- O A few times
- O Never

**33. DURING THE PAST 12 MONTHS, how often were pesticides used INSIDE your residence to control for insects?** *If the frequency changed throughout the year, report the highest frequency.*
- O More than once a week
- O Once a week
- O Once a month
- O Once every 2-5 months
- O Once every 6 months
- O Once during the past 12 months
- O Never
- O Don’t know

**34. DURING THE PAST 12 MONTHS, how often were pesticides used OUTSIDE your residence to control for insects?** *If the frequency changed throughout the year, report the highest frequency.*

- O More than once a week
- O Once a week
- O Once a month
- O Once every 2-5 months
- O Once every 6 months
- O Once during the past 12 months
- O Never
- O Don't know

**35. In the past month, have you noticed any unusual or persistent odors in your house?**

- O Yes
- O No

**36. Have you or any household members experienced any symptoms while indoors in the past month?**

- O Yes
- O No

If so, please describe the symptoms:

______________________________________________________________________

______________________________________________________________________

**37. Please provide any additional information or concerns you have about the air quality in your house.**

______________________________________________________________________

______________________________________________________________________

______________________________________________________________________

**OK-AIR Study**

**Parent Questionnaire – Baseline Health Assessment**

**Study ID: ______________________________**

**About the person completing the questionnaire:**

**1. My name: __________________________________**

**2. I am the:**
- ○ Child's mother
- ○ Child's father
- ○ Female guardian
- ○ Male guardian
- ○ Other specific relation: ___________________________

**About your child:**

**3. Child's name:**

First name: ____________________
Middle name: _________________
Last name: _______________________

**4. Child's date of birth: __/__/____**

**5. Was this child born prematurely?**
- ○ No
- ○ Yes
- ○ I do not know

**6. At what age did your child first start group childcare such as daycare or Head Start?**
- ○ As an infant under age 1
- ○ Between age 1 and 2
- ○ Between age 2 and 3
- ○ At age 3
- ○ At age 4

**7. Did your child first start group care within the last three months?**
- ○ Yes
- ○ No

**8. What was your child's biological sex at birth (circle one):**
- ○ Male
- ○ Female
- ○ Other: ___________
- ○ I prefer not to say

**9. Is your child of Hispanic, Latino, or Spanish origin?**

- ○ Yes
- ○ No

**10. What is your child's race(s)?** *Please mark ALL that apply.*

- ❑ American Indian or Alaska Native
- ❑ Asian
- ❑ Black or African American
- ❑ Native Hawaiian or other Pacific Islander
- ❑ White or Caucasian
- ❑ Other (please specify): ______________
- ❑ I prefer not to say

**11. In general, would you say your child's overall health is:**

- ❑ excellent
- ❑ very good
- ❑ good
- ❑ fair
- ❑ poor

**CHILDREN'S ACTIVITIES**

**12. How much time would your child usually spend watching television, using a computer or other screen:**

**a. On a <u>weekend</u> day:**

- ❑ less than one hour each day
- ❑ 1-2 hours each day
- ❑ 3 or more hours each day

**b. On a <u>school</u> day:**

- ❑ less than one hour each day
- ❑ 1-2 hours each day
- ❑ 3 or more hours each day

**13. How much time would your child usually spend playing outdoors (alone or with other children):**

| **In spring or fall:** | **In summer:** | **In winter:** |
|---|---|---|
| AFTER SCHOOL<br>○ _____ hours _____minutes<br><br>ON WEEKENDS/VACATION DAYS<br>○ _____ hours _____minutes | AFTER SCHOOL<br>○ _____ hours _____minutes<br><br>ON WEEKENDS/VACATION DAYS<br>○ _____ hours _____minutes | AFTER SCHOOL<br>○ _____ hours _____minutes<br><br>ON WEEKENDS/VACATION DAYS<br>○ _____ hours _____minutes |

**14. In the past 12 months, how often have problems with transportation interfered with your ability to get to school, to work, to medical appointments or to meet family responsibilities?**

- ○ Never
- ○ Sometimes
- ○ Usually
- ○ Always

**QUESTIONS ABOUT ASTHMA**

**15. Has your child ever had wheezing or whistling in the chest at any time in the past?**

- O Yes
- O No

*IF YOU HAVE ANSWERED "NO" PLEASE SKIP TO QUESTION 28*

**16. Has your child had wheezing or whistling in the chest in the past 12 months?**

- O Yes
- O No

*IF YOU HAVE ANSWERED "NO" PLEASE SKIP TO QUESTION 28*

**17. How many attacks of wheezing has your child had in the past 12 months?**

- O None
- O 1 to 3
- O 4 to 12
- O More than 12

**18. In the past 12 months, how often, on average, has your child's sleep been disturbed due to wheezing?**

- O Never woken with wheezing
- O Less than one night per week
- O One or more nights per week

**19. In the past 12 months, has wheezing ever been severe enough to limit your child's speech to only one or two words at a time between breaths?**

- O Yes
- O No

**20. Has a doctor told you that your child has asthma?**

- O Yes
- O No

**21. In the past 12 months, has your child's chest sounded wheezy during or after exercise?**

- O Yes
- O No

**22. In the past 12 months, has your child had a dry cough at night, apart from a cough associated with a cold or chest infection?**

- O Yes
- O No

**ASTHMA CONTROL**
(If you answered no to Question 15 or Question 16, skip this section.)

**23. During the past 4 weeks, how often was your child bothered by breathing problems, such as wheezing, coughing, or shortness of breath?**
- Not at all
- Once or twice
- Once every week
- 2 or 3 times per week
- 4+ times per week

**24. During the past 4 weeks, how often did your child's breathing problems (wheezing, coughing, shortness of breath) wake him or her up at night?**
- Not at all
- Once or twice
- Once every week
- 2 or 3 times per week
- 4+ times per week

**25. During the past 4 weeks, to what extent did your child's breathing problems, such as wheezing, coughing, or shortness of breath, interfere with his or her ability to play, go to school, or engage in usual activities that a child should be doing at his or her age?**
- Not at all
- Slightly
- Moderately
- Quite a lot
- Extremely

**26. During the past 3 months, how often did you need to treat your child's breathing problems** (wheezing, coughing, shortness of breath) **with quick-relief medications** (albuterol, Ventolin©, Proventil©, Maxair©, ProAir©, Xopenex©, or Primatene© Mist)?
- Not at all
- Once or twice
- Once every week
- 2 or 3 times per week
- 4+ times per week

**27. During the past 12 months, how often did your child need to take oral corticosteroids** (prednisone, prednisilone, Orapred©, Prelone©, or Decadron©) **for breathing problems not controlled by other medications?**
- Never
- Once
- Twice
- 3 times
- 4+ times

**QUESTIONS ABOUT CHRONIC RESPIRATORY SYMPTOMS**

**28. Has your child ever had a problem with sneezing, or a runny, or blocked nose when he/she DID NOT have a cold or the flu?**

- O Yes
- O No

*IF YOU HAVE ANSWERED "NO" PLEASE SKIP TO QUESTION 34*

---

**29. In the past 12 months, has your child had a problem with sneezing, or a runny, or blocked nose when he/she DID NOT have a cold or the flu?**

- O Yes
- O No

*IF YOU HAVE ANSWERED "NO" PLEASE SKIP TO QUESTION 34*

---

**30. In the past 12 months, has this nose problem been accompanied by itchy-watery eyes?**

- O Yes
- O No

**31. In which of the past months did this nose problem occur?** *Please mark all that apply.*

- O Fall (Sept-Nov)
- O Winter (Dec-Feb)
- O Spring (Mar-May)
- O Summer (June-Aug)

**32. In the past 12 months, how much did this nose problem interfere with your child's daily activities?**

- O Not at all
- O A little
- O A moderate amount
- O A lot

**33. Has your child ever had hay fever?**

- O Yes
- O No

**QUESTIONS ABOUT ECZEMA**

**34. Has your child ever had an itchy rash which was coming and going for at least six months?**

- O Yes
- O No

*IF YOU HAVE ANSWERED "NO" PLEASE SKIP TO QUESTION 36*

---

**35. Has your child had this itchy rash at any time in the past 12 months?**

- O Yes
- O No

**RESIDENTIAL HISTORY**

**36. What is the address where your child lives with you, currently?**
STREET ADDRESS:_________________________________
CITY:_______________________
STATE:________
ZIP:_____________

**37. How long have you lived together with your child at your current address?**

__ __ Years __ __ Months (*If less than 12 months*)

**HOUSING**

**38. What is the type of housing where you currently live with your child?**
- O Apartment
- O Duplex or townhouse
- O Single family home
- O Mobile home
- O Temporary housing (for example, women's shelter, homeless shelter)
- O Motel/ hotel
- O Boat, RV, or van
- O Other (describe): __________________________

(a) **This house, apartment, or mobile home is:**
- O Owned by you or someone in this household with a mortgage or loan (include home equity loans)
- O Owned by you or someone in this household free and clear (without a mortgage or loan)
- O Rented
- O Occupied without payment of rent
- O Not applicable
- O Other (describe): _________________________

(b) **How much do you spend on your rent or mortgage per month?** $__________

(c) **How many rooms are there in your current home?** Please include all rooms such as the kitchen and living room, but not bathrooms or hallways. _________

(d) **What is the approximate size of your house in square feet?** _______ sq. ft.

**(e) How many people live in your current home?**

________ adults
________ children (aged 0-18 y)

Age of the youngest person living in your home: ________ years __________ months

**(f) In your current home, how many people share a bedroom with your child?**
- O the child has his / her own bedroom
- O the child shares a bedroom with 1 person
- O the child shares a bedroom with 2 people
- O the child shares a bedroom with more than 2 people

**(g) How is your current home heated?**

- ○ Built-in electric units
- ○ Warm air furnace (central heating unit run on gas, propane, etc.)
- ○ Floor, wall, or pipeless furnace (does not use ducts to move warm air throughout the home)
- ○ Steam or hot water
- ○ Other means: ________________________
- ○ Not heated
- ○ I do not know

**(h) What fuel is used most for cooking in your home?**

- ○ Utility gas
- ○ Electricity
- ○ Bottled, tank or LP gas
- ○ Wood
- ○ Fuel oil, kerosene
- ○ Coal or coke

**(i) Do you use the following equipment in your home?** *Check all that apply.*

- ❑ Central air conditioning (HVAC)
- ❑ Air conditioning window / wall unit(s)
- ❑ Humidifiers(s)
- ❑ Air filtration unit
- ❑ None of these

**(j) Do you use any of the following regularly in your home?** *Check all that apply.*

- ❑ Air fresheners or sprays
- ❑ Scented candles
- ❑ Incense
- ❑ None of these

**(k) Do you have pets living indoors in your home?** *Check all that apply.*

- ❑ No
- ❑ Yes, cat(s)
- ❑ Yes, dog(s)
- ❑ Yes, bird(s)
- ❑ Yes, other: ________________________

**(l) How often do you open windows in your home for ventilation?**

| **In spring:** | **In summer:** | **In fall:** | **In winter:** |
|---|---|---|---|
| ○ Every day<br>○ At least once most weeks<br>○ Occasionally<br>○ Rarely | ○ Every day<br>○ At least once most weeks<br>○ Occasionally<br>○ Rarely | ○ Every day<br>○ At least once most weeks<br>○ Occasionally<br>○ Rarely | ○ Every day<br>○ At least once most weeks<br>○ Occasionally<br>○ Rarely |

### HOUSING QUALITY

**39. Overall, how would you describe the physical [or structural] condition of your current home?**

- ○ Excellent
- ○ Good
- ○ Fair
- ○ Poor

**40. Now I am going to ask you some questions about problems that people have in some homes or apartments. How much of a problem is…**

**a. A heating system that does not work?**

- ○ Big Problem
- ○ Small Problem
- ○ No Problem at all

**b. Flooding or water infiltration?**

- ○ Big Problem
- ○ Small Problem
- ○ No Problem at all

**c. Mold or mildew?**

- ○ Big Problem
- ○ Small Problem
- ○ No Problem at all

### ENVIRONMENTAL HEALTH

**41. In the last 12 months, how often has anyone, including visitors, smoked tobacco inside your home?**

- ○ Daily
- ○ Weekly
- ○ Monthly
- ○ A few times
- ○ Never

**42. In the last 12 months, how often has tobacco smoke entered your home from somewhere else in or around the building?**

- ○ Daily
- ○ Weekly
- ○ Monthly
- ○ A few times
- ○ Never

**43. DURING THE PAST 12 MONTHS, how often were pesticides used INSIDE your residence to control for insects?** *If the frequency changed throughout the year, report the highest frequency.*

- ○ More than once a week
- ○ Once a week
- ○ Once a month
- ○ Once every 2-5 months
- ○ Once every 6 months
- ○ Once during the past 12 months
- ○ Never
- ○ Don't know

**44. DURING THE PAST 12 MONTHS, how often were pesticides used OUTSIDE your residence to control for insects?** *If the frequency changed throughout the year, report the highest frequency.*

- ○ More than once a week
- ○ Once a week
- ○ Once a month
- ○ Once every 2-5 months
- ○ Once every 6 months
- ○ Once during the past 12 months
- ○ Never
- ○ Don't know

**45. In the past month, have you noticed any unusual or persistent odors in your house?**

- ○ Yes
- ○ No

**46. Have you or any household members experienced any symptoms while indoors in the past month?**

- ○ Yes
- ○ No

If so, please describe the symptoms:

________________________________________

________________________________________

**47. Please provide any additional information or concerns you have about the air quality in your house.**

________________________________________

________________________________________

________________________________________

________________________________________

________________________________________
